# Prevalence and work-related risk factors of Musculoskeletal Disorders among miners at Kalumbila Mine, Kalumbila District Zambia

**DOI:** 10.1101/2024.05.15.24307432

**Authors:** Chibuye Kunda, Joseph Lupenga, Chisala D. Meki

## Abstract

**Background:** Mining is a hazardous occupation with high injury rates and work-related musculoskeletal disorders. However, few studies have reported on the prevalence and risk factors of work related musculoskeletal disorders among mineworkers in Zambia. Therefore, the study sought to examine risk factors of work-related musculoskeletal disorders among mineworkers at Kalumbila mine in Zambia.

**Method:** A cross-sectional study was undertaken and data were collected using a selfadministered Cornell Musculoskeletal Discomfort Questionnaire. A total of 357 participants were selected using a systematic random sampling method from among the male and female mine workers at the Kalumbila mine. Stata 17 was used to analyse the data and the descriptive analysis was used to determine the prevalence and types of work-related musculoskeletal disorders. Factors associated with work-related musculoskeletal disorders were identified using simple and multiple logistic regression. The level of significance was set at 0.05 and confidence level at 95%.

**Results:** The study revealed 274/356 (77%) of the participants reporting pain or discomfort in at least one body location. The highest level of discomfort or pain was reported in the lower back 184 (51.5%), upper back 90 (25.2%) and neck 89 (24.9%), whereas the lowest level of discomfort was reported in the left thigh 10 (2.8%), left forearm 11 (3.1%) and left foot 13 (3.6%). In the multivariable analysis, secondary education (AOR 2.03 CI 95%: 1.02 - 4.05, p=0.044), not taking any breaks while working (AOR 0.10 CI 95%: 0.01 - 0.86, p=0.036), and operating machinery causing whole-body vibration (AOR 3.0 CI 95%: 1.52-5.95, p=0.002) were associated with work-related musculoskeletal disorders.

**Conclusion:** Self-reported work-related musculoskeletal disorders were common among miners, underscoring the need to invest in worker safety through ergonomic programs and workers’ training on safety measures.

## Introduction

Since the establishment of Zambia as a nation, copper has been the single largest contributor to the Zambian economy. At independence in 1964, copper accounted for 91% of the total export earnings. In 2016, mining accounted for 12% of Zambiás GDP and 70% of total export value (1). The sector is a significant source of government revenue and employment and hence remains critically important for the Zambian economy (2).

Mining is characterised by most researchers as one of the most hazardous occupations amongst major industrial activities (3). Mining activities involves drilling, charging and blasting to access and recover ore and mineworkers are often required to perform labour-intensive tasks that expose them to greater occupational hazards. The mining sector is associated with greater injury rates than other sectors and mine employees are often subjected to a higher danger of work-related musculoskeletal disorders (WMSD) (4).

Work-related musculoskeletal disorders (WMSDs) are a common type of occupational injuries worldwide (5). There is compelling evidence that work-related musculoskeletal disorders (WMSDs) affect mineworkers to a greater degree than workers in other industries (5). For example, studies have shown that miners experience more disability from knee and back pain, more absenteeism, more osteoarthritis, and more disk degeneration compared to other industrial populations (6).

According to the International Labour Organization (ILO), among the approximately 160 million work related illnesses that occur worldwide each year, work related musculoskeletal disorders (WMSDs) are known to be the second most common occupational disease (3).

Despite regulations, automation and increased attention towards reducing risks through safety campaigns, the mining industry is still associated with higher rates of injuries compared with other industries (5). Mineworkers must deal with a number of subtly harmful risks to safety and health, such as a high concentration of mechanical equipment in a confined space (7).

Most mining companies in developing countries such as Zambia do not have data regarding the prevalence of WMSDs. To date, only a few studies have reported on the prevalence and risk factors associated with WMSDs amongst mineworkers in Zambia.

Therefore, a description and understanding of the effects of exposure profiles in mining based on the results of epidemiological investigations is required (8). It is evident from the facts summarized above that WMSDs in miners is a critical area of concern that must be addressed, regularly from an ergonomics perspective.

The study sought to examine risk factors that are associated with work-related musculoskeletal disorders (WMSDs) among mineworkers at Kalumbila mine in Zambia.

## Materials and methods

### Study design and setting

This study adopted a cross-sectional research design to assess the MSDs and their associated factors among the mineworkers of Kalumbila Mine in the North-western province of Zambia.

A quantitative method was used to assess the risk factors that contribute to the MSDs. First Quantum is a global copper company. With production of copper in the form of concentrate, cathode and anode, and have inventories of nickel, gold and cobalt. The Kalumbila open-pit copper mine, 150km west of Solwezi in North Western Province of Zambia, is at the forefront of mining technology.

### Study population and sample size

The study was conducted among mineworkers at Kalumbila mine, the mine has 3362 employees spread across departments including Human resource, Mining Operations, Mining Technical, Health and safety, Engineering, Process plant, Commercial and Site services. Using the sample size formula by Yamane (1967) for cross-sectional study:

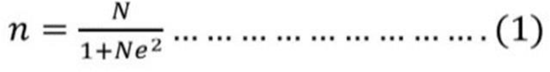

Sample size of 356, formula by Yamane was used to calculate sample size due to an infinite population.

### Data source

To meet the required sample size of 356 and ensure proportionate representation of the population across the mine, a stratified random sampling method was used according to departments across the mine sites. This study’s data was collected using a questionnaire comprising of four sections.

The mineworkers who fulfilled the inclusion criteria were selected as the respondents of this study. These included those who are current employees at the mine, those who are 18 years and above, with minimum of one-year experience at the mine and signed or given informed consent to participate in the study.

Excluded were those who had worked for less than a year at the mine, those who are not employees of Kalumbila Mine and refusal to consent to participate in the study.

Various questionnaires guided the design of the questionnaire for this study including the standardized Nordic, and the modified versions of the Washington state risk factor checklist used by (6).

The Nordic questionnaire is a widely accepted, easy to administer, and costefficient tool for collecting data on selfreported musculoskeletal discomfort and sickness, it has been shown to have high validity for capturing MSDs in various settings (9).

### Ethical Issues

Ethical issues involved in the study were addressed by doing the following. Ethics approval was obtained from the Ethics Review Committee through the School of Public Health at The University of Zambia. Consent was also obtained from the National Health Research Authority as clearance to carry out research. Approval was secured after a letter of introduction from the School of Public Health (SPH), University of Zambia to Kalumbila Minerals Limited for approval of data collection from the various mine departments and allow easy access to information needed for the study.

Consent was sought from workers prior to participating in the study. The researcher did not anticipate any potential risks of participation to participants. Most of the questions were not sensitive to inflict any emotional injury on participants. No names of participants were indicated on the questionnaires.

Participants were assured of confidentiality and privacy of the information provided. Information was gathered with a structured questionnaire. The research instrument (questionnaire) containing the data was saved under lock and key accessible only to the principal investigator.

### Variables and measurements

Our dependent variable was work-related musculoskeletal disorders coded as 1 if symptoms of pain in the back, neck, shoulder, and knee and wrist were present in the last 12 months and 2 if otherwise. The independent variables were grouped into individual or personal factors, physical factors and psychological factors.

Individual factors include: Age (>20, 2030,30-40, 50-60, 60 and above); Sex (Male, Female); Marital status (Married or others); Level of education (no formal education, primary education, secondary education, trade school, tertiary education); Smoking (yes or no); Department, position and length of service.

Physical factors included: Working schedule (days only, day and night); number of hours worked per day (<8 hours per day, >8 hours per day); breaks in a day (none, once, twice, more than 3 times); Body vibrations (Yes, no); Lifting >25kgs (none, 1-5 times, >5 times); Standing work (less than 1 hour, 1-4 hrs., 5-8hrs, more than 9 hrs.); working overhead (less than 1 hour, 1-4 hours, 5-8 hours, >9 hours).

Psychological factors included: Indicating the degree to which they agree with strongly disagree (1), disagree (2), agree (3), strongly agree (4) to the following statements; my job requires that I learn new things; my job involves a lot of repetitive work; my job requires a high level of skill; my job requires working fast; my job requires working very hard; my supervisor pays attention to what I am saying.

### Analysis

The analysis starts with descriptive statistics namely frequencies expressed as percentages on the prevalence of MSDs and affected anatomical sites. The univariate (unadjusted) and multivariable (adjusted) logistic regression analysis was performed to assess the association between the independent variables and the outcome variable. WMSD reporting for odds ratios (ORs) and 95% confidence interval (95 % CI).

Variables with a p<0.25 in the univariate analysis and those that were clinically important were included in the multivariate analysis (Chowdhury & Turin, 2020). To create the final model, backward elimination logistic regression was used retaining all explanatory variables with a pvalue <0.20.

In model selection, the Bayesian information criterion (BIC) and Akaike’s information criterion (AIC) were used, with the model with the lowest value being preferred. The Hosmer–Lemeshow test was used to evaluate the goodness of fit of the final models.

### Ethical considerations

Ethics approval was obtained from the Ethics Review Committee through the School of Public Health at The University of Zambia and the National Health Research Authority as clearance to carry out research.

Data was collected between 14^th^ September 2023 and 20^th^ December 2023. Written consent was obtained from the participants prior to them responding to the questionnaires or participating in the study.

## Results

### Demographic and Work-related Characteristics

Approximately forty-four percent (159/356) of the participants were aged between 30 and 40 years, while 105 (29.5%) of the respondents were aged between 41 and 50 years. Males made up the majority of the participants in the study 295 (83.8 %).

Majority of the respondents-149 (41.9%) were from the Mining Department, 97 (27.2%) were from the Maintenance department, 59 (16.6%) from Engineering, process plant had 27 (7.6%)while 24 (6.7%) were from other departments.

Most participants were married 266 (74.7%), and in terms of educational level, 145 (40.7%) had a tertiary education and 139 (39.0%) had attended a trade school. Most of the participants were not current smokers 295 (83.6%) (Table 1).

**Table 1:** Demographic characteristics of the study participants from Kalumbila Mine.

| Demographic Characteristics | Frequency | Percentage |
| --- | --- | --- |
| Age<br>20 – 30 | 45 | 12.6% |
| 30 – 40 | 159 | 44.7% |
| 40 – 50 | 105 | 29.5% |
| 50 – 60 | 33 | 9.3% |
| 60 and above | 14 | 3.9% |
| Total | 356 | 100.0% |
| **Sex** |  |  |
| Female | 57 | 16.2% |
| Male | 295 | 83.8% |
| Missing data | 4 |  |
| Total | 356 | 100.0% |
| **Marital Status** Divorced |  |  |
|  | 19 | 5.3% |
| Married | 266 | 74.7% |
| Single | 67 | 18.8% |
| Widowed | 4 | 1.1% |
| Total | 356 | 100.0% |
| **Educational Level** No formal education |  |  |
|  | 6 | 1.7% |
| Primary School | 1 | 0.3% |
| Secondary School | 65 | 18.3% |
| Tertiary School | 145 | 40.7% |
| Trade School | 139 | 39.0% |
| Total | 356 | 100.0% |
| **Smoking** |  |  |
| No | 295 | 83.6% |
| Yes | 58 | 16.4% |
| Missing data | 3 |  |
| Total | 356 | 100.0% |
| **Body Mass Index** Underweight |  |  |
|  | 1 | 29.0% |
| Healthy weight | 135 | 38.8% |
| Overweight | 149 | 42.8% |
| Obese | 63 | 18.1% |
| Missing data | 7 |  |
| Total | 356 | 100.0% |
| Median Length of service (interquartile range) | 5 (3 - 7) |  |

77% of the participants reported pain or discomfort in at least one body location (Figure 1). The highest level of discomfort or pain was reported in the lower back 184 (51.5%), upper back 90 (25.2%) and neck 89 (24.9%), whereas the lowest level of discomfort was reported in the left thigh 10 (2.8%), left forearm 11 (3.1%) and left foot 13 (3.6%).

**Figure 1:**
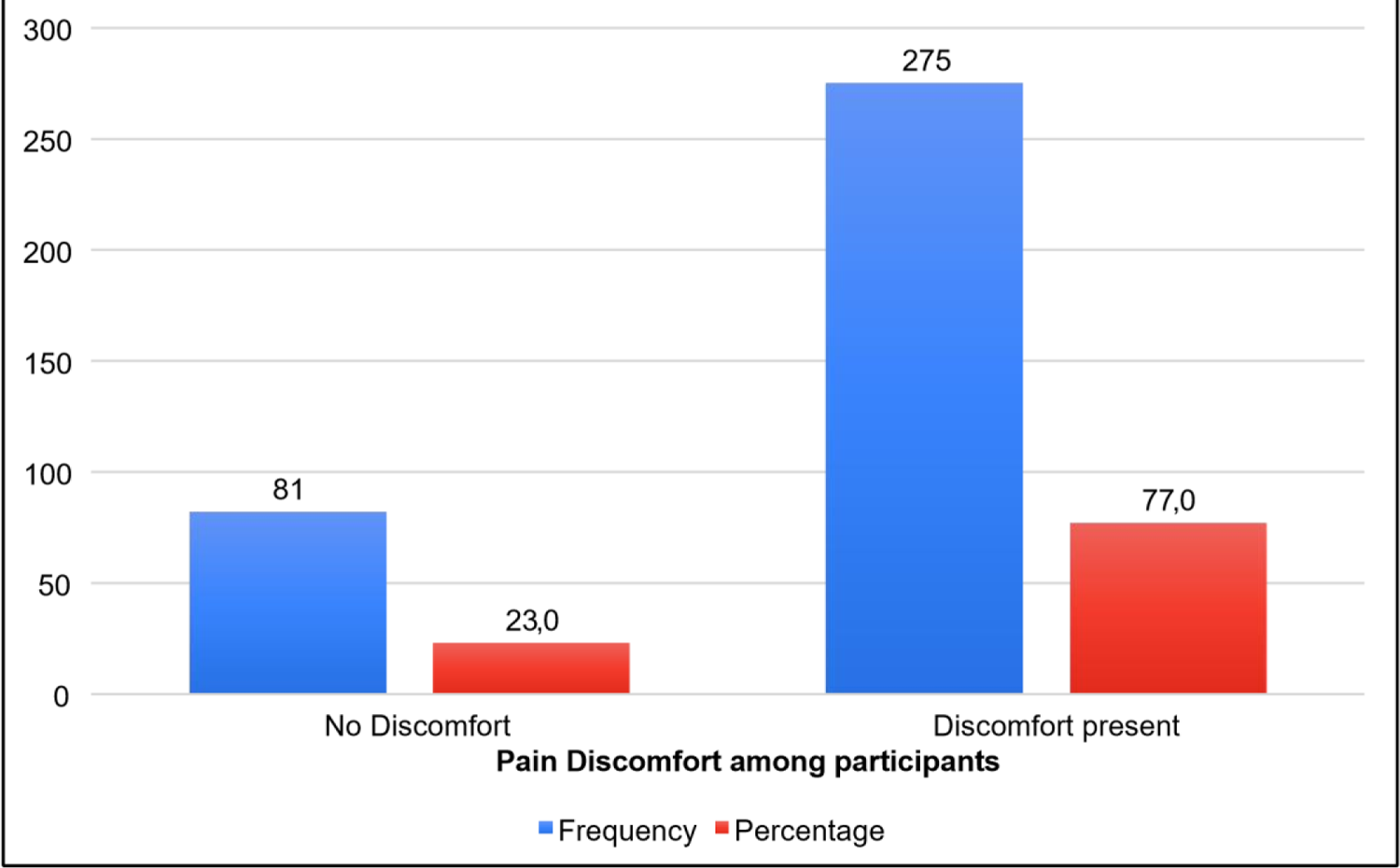
Discomfort in at least one body location

Table 2 shows the prevalence of musculoskeletal disorders in different anatomical regions of the study participants.

**Table 2:**
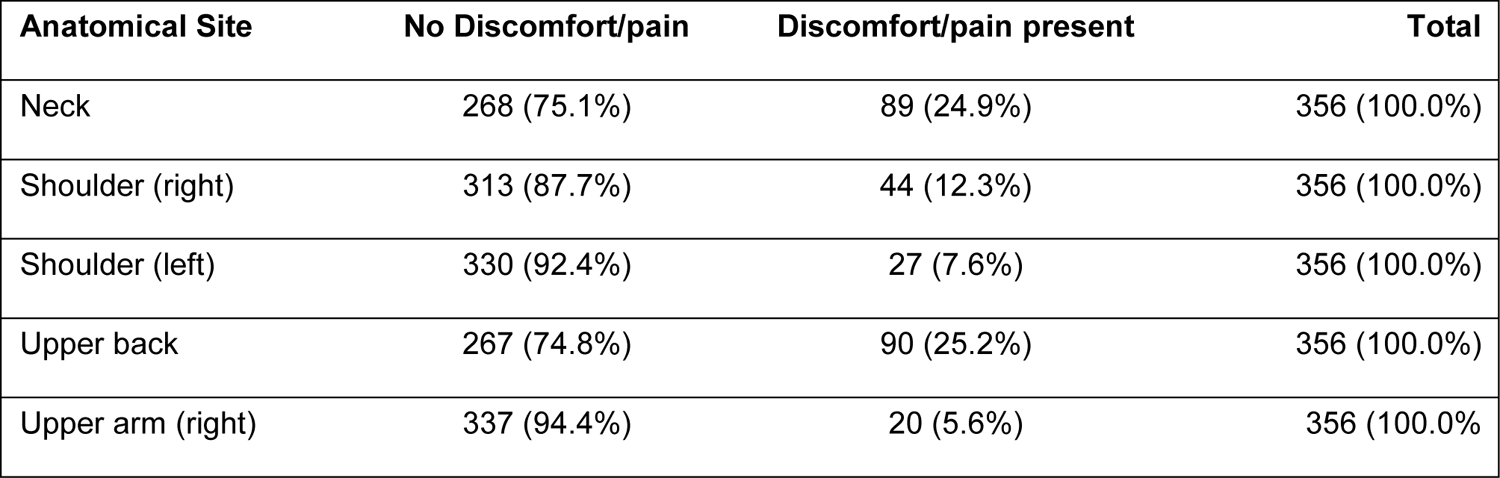
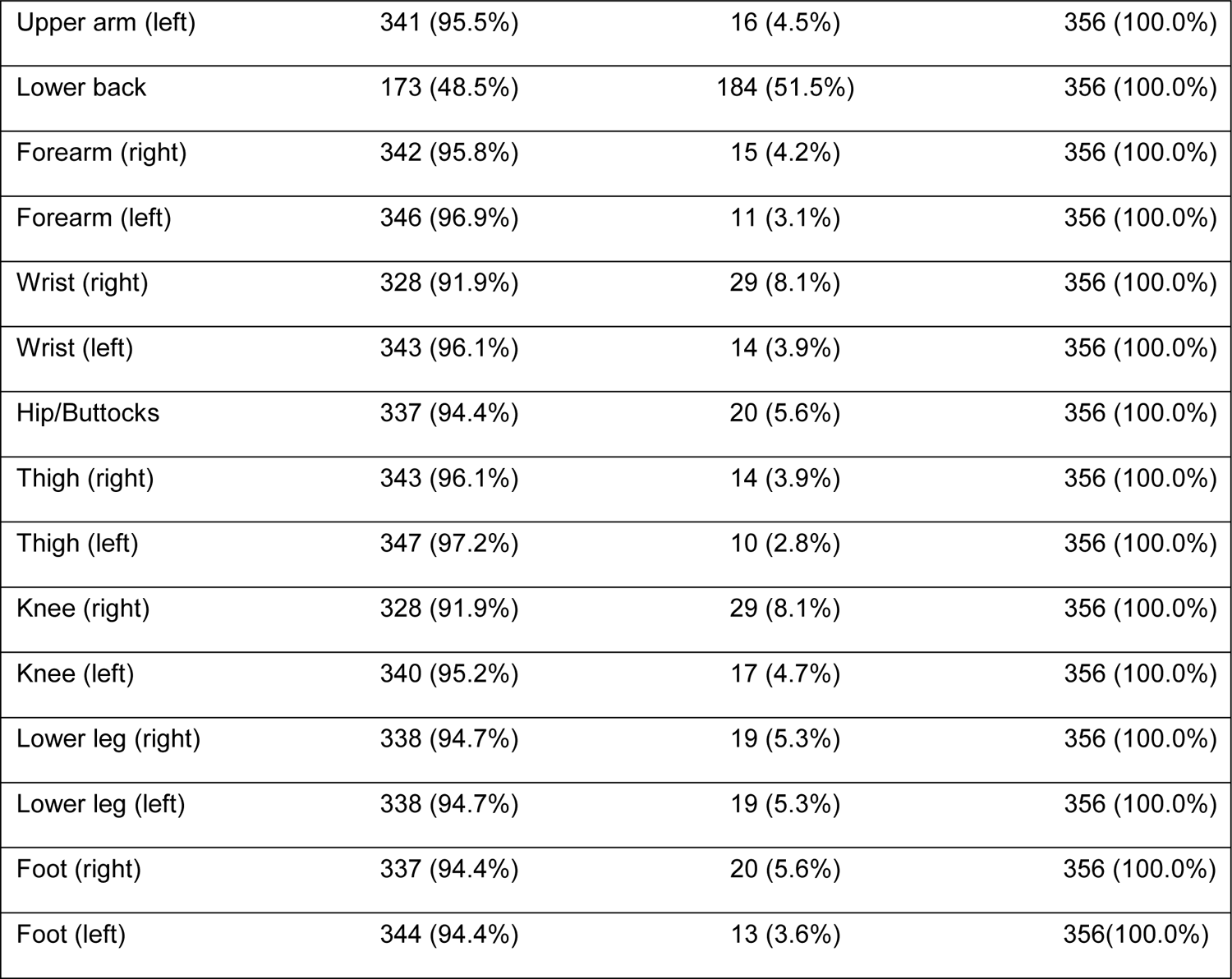
Prevalence of musculoskeletal disorders in different anatomical regions of the study participants (n=357).

| Anatomical Site | No Discomfort/pain | Discomfort/pain present | Total |
| --- | --- | --- | --- |
| Neck | 268 (75.1%) | 89 (24.9%) | 356 (100.0%) |
| Shoulder (right) | 313 (87.7%) | 44 (12.3%) | 356 (100.0%) |
| Shoulder (left) | 330 (92.4%) | 27 (7.6%) | 356 (100.0%) |
| Upper back | 267 (74.8%) | 90 (25.2%) | 356 (100.0%) |
| Upper arm (right) | 337 (94.4%) | 20 (5.6%) | 356 (100.0%) |
| Upper arm (left) | 341 (95.5%) | 16 (4.5%) | 356 (100.0%) |
| Lower back | 173 (48.5%) | 184 (51.5%) | 356 (100.0%) |
| Forearm (right) | 342 (95.8%) | 15 (4.2%) | 356 (100.0%) |
| Forearm (left) | 346 (96.9%) | 11 (3.1%) | 356 (100.0%) |
| Wrist (right) | 328 (91.9%) | 29 (8.1%) | 356 (100.0%) |
| Wrist (left) | 343 (96.1%) | 14 (3.9%) | 356 (100.0%) |
| Hip/Buttocks | 337 (94.4%) | 20 (5.6%) | 356 (100.0%) |
| Thigh (right) | 343 (96.1%) | 14 (3.9%) | 356 (100.0%) |
| Thigh (left) | 347 (97.2%) | 10 (2.8%) | 356 (100.0%) |
| Knee (right) | 328 (91.9%) | 29 (8.1%) | 356 (100.0%) |
| Knee (left) | 340 (95.2%) | 17 (4.7%) | 356 (100.0%) |
| Lower leg (right) | 338 (94.7%) | 19 (5.3%) | 356 (100.0%) |
| Lower leg (left) | 338 (94.7%) | 19 (5.3%) | 356 (100.0%) |
| Foot (right) | 337 (94.4%) | 20 (5.6%) | 356 (100.0%) |
| Foot (left) | 344 (94.4%) | 13 (3.6%) | 356(100.0%) |

### Psychological factors

Most participants agreed that their job required them to learn new things 305 (87.0%), involved a lot of repetitive work 302 (87.5%), required them to be creative 331 (96.5%), allowed them to make a lot of decisions 213 (62.1%) and required a high level of skills 324 (94.5%).

In addition, most participants agreed that they were allowed to decide how to do the job 206 (59.9%), that they get to do a variety of things at work 310 (90.4%), and that they had a lot to say about what happens at work 236 (68.8%). Participants also agreed that their job required them to work very fast 298 (87.6%), required them to work very hard 321 (94.4%), that they had enough time to get the job done 308 (90.3%), that their colleagues helped them when the job got difficult 324 (94.5%) and that their supervisors paid attention to what they said 321 (93.0%) (Table 3).

**Table 3:**
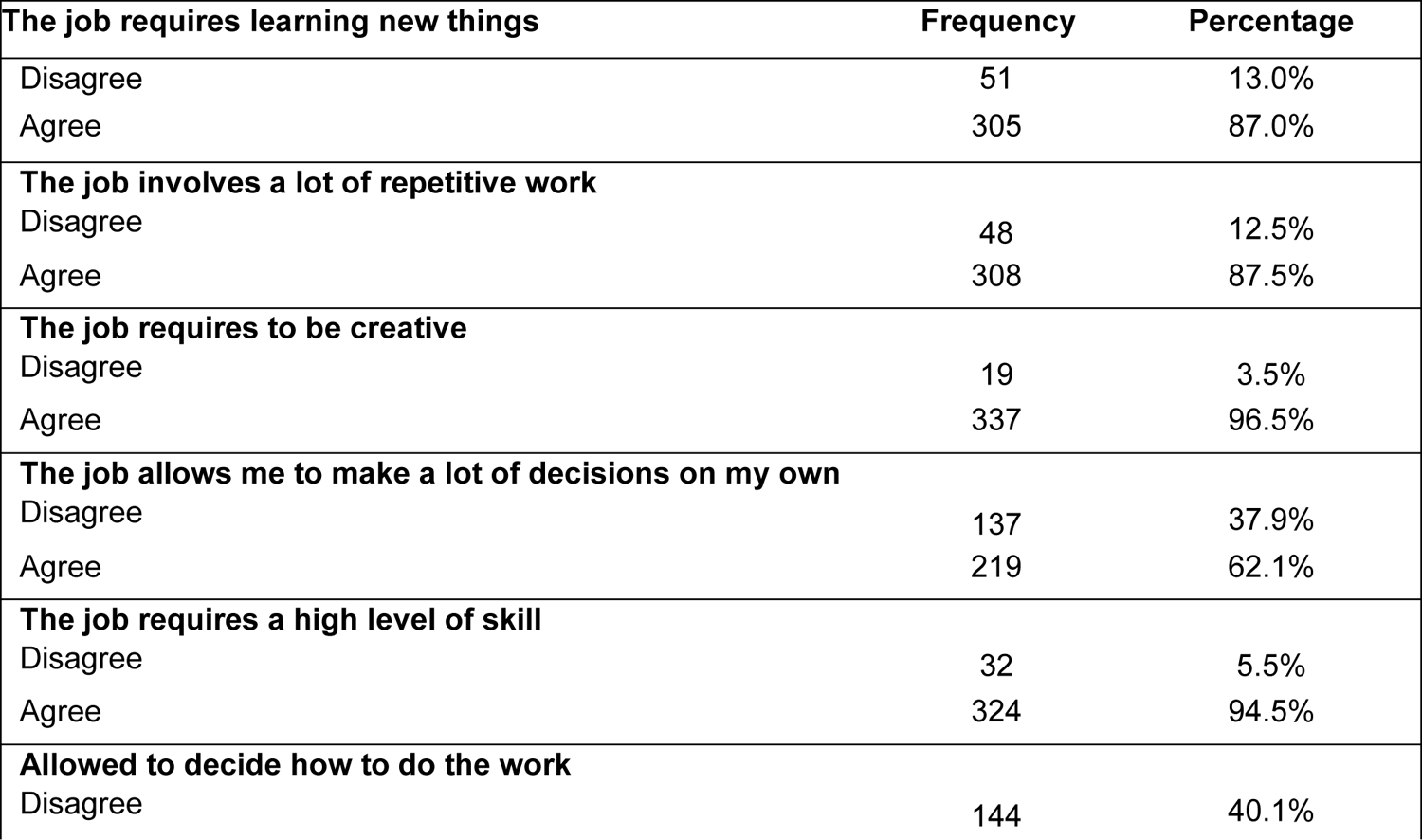
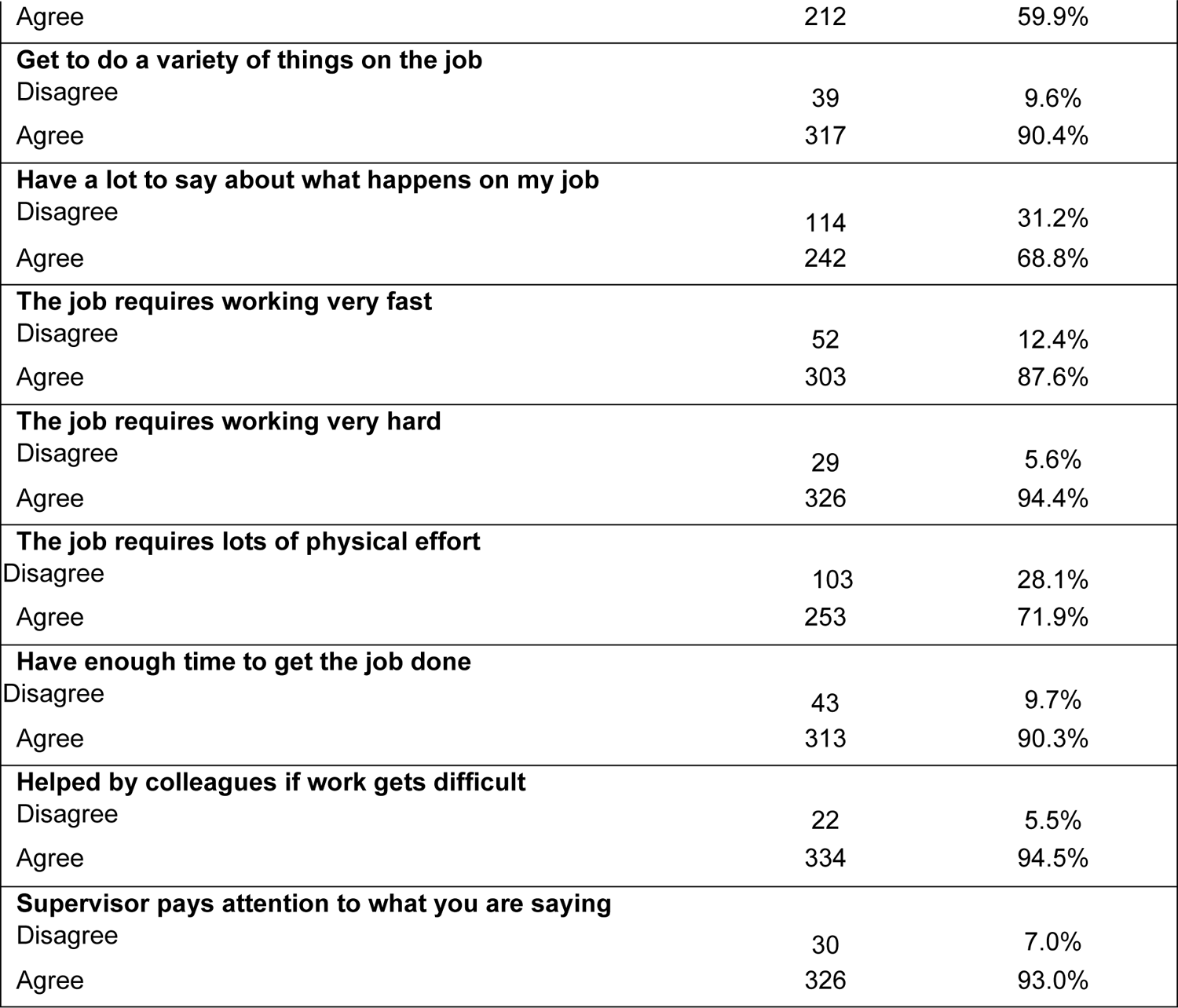
Work-related information about the study participants.

| **The job requires learning new things** | **Frequency** | **Percentage** |
| --- | --- | --- |
| Disagree | 51 | 13.0% |
| Agree | 305 | 87.0% |
| **The job involves a lot of repetitive work** |  |  |
| Disagree | 48 | 12.5% |
| Agree | 308 | 87.5% |
| **The job requires to be creative** |  |  |
| Disagree | 19 | 3.5% |
| Agree | 337 | 96.5% |
| **The job allows me to make a lot of decisions on my own** |  |  |
| Disagree | 137 | 37.9% |
| Agree | 219 | 62.1% |
| **The job requires a high level of skill** |  |  |
| Disagree | 32 | 5.5% |
| Agree | 324 | 94.5% |
| **Allowed to decide how to do the work** |  |  |
| Disagree | 144 | 40.1% |
| Agree | 212 | 59.9% |
| **Get to do a variety of things on the job** |  |  |
| Disagree | 39 | 9.6% |
| Agree | 317 | 90.4% |
| **Have a lot to say about what happens on my job** |  |  |
| Disagree | 114 | 31.2% |
| Agree | 242 | 68.8% |
| **The job requires working very fast** |  |  |
| Disagree | 52 | 12.4% |
| Agree | 303 | 87.6% |
| **The job requires working very hard** |  |  |
| Disagree | 29 | 5.6% |
| Agree | 326 | 94.4% |
| **The job requires lots of physical effort** |  |  |
| Disagree | 103 | 28.1% |
| Agree | 253 | 71.9% |
| **Have enough time to get the job done** |  |  |
| Disagree | 43 | 9.7% |
| Agree | 313 | 90.3% |
| **Helped by colleagues if work gets difficult** |  |  |
| Disagree | 22 | 5.5% |
| Agree | 334 | 94.5% |
| **Supervisor pays attention to what you are saying** |  |  |
| Disagree | 30 | 7.0% |
| Agree | 326 | 93.0% |

### Factors associated with WMSDs among mineworkers

Both univariate and multiple logistic regression analyses were performed. In the univariate analysis, age, educational level, working long hours while standing and having a job that requires high skills were associated with WMSD among mine workers (p<0.05).

In the multivariable analysis, those with secondary education were 2.03 times more likely to have WMSD than those with vocational education (AOR 2.03 CI 95%: 1.02 - 4.05, p=0.044).

Those who did not take any breaks while working were 0.30 times less likely to experience WMSD compared with those who took more than 2 breaks in a typical working day (AOR 0.10 CI 95%: 0.01 - 0.86, p=0.036).

Participants who operated machinery causing whole-body vibration were 3.0 times more likely to experience WMSD than those who did not (AOR 3.0 CI 95%: 1.52-5.95, p=0.002) (Table 4)

**Table 4:**
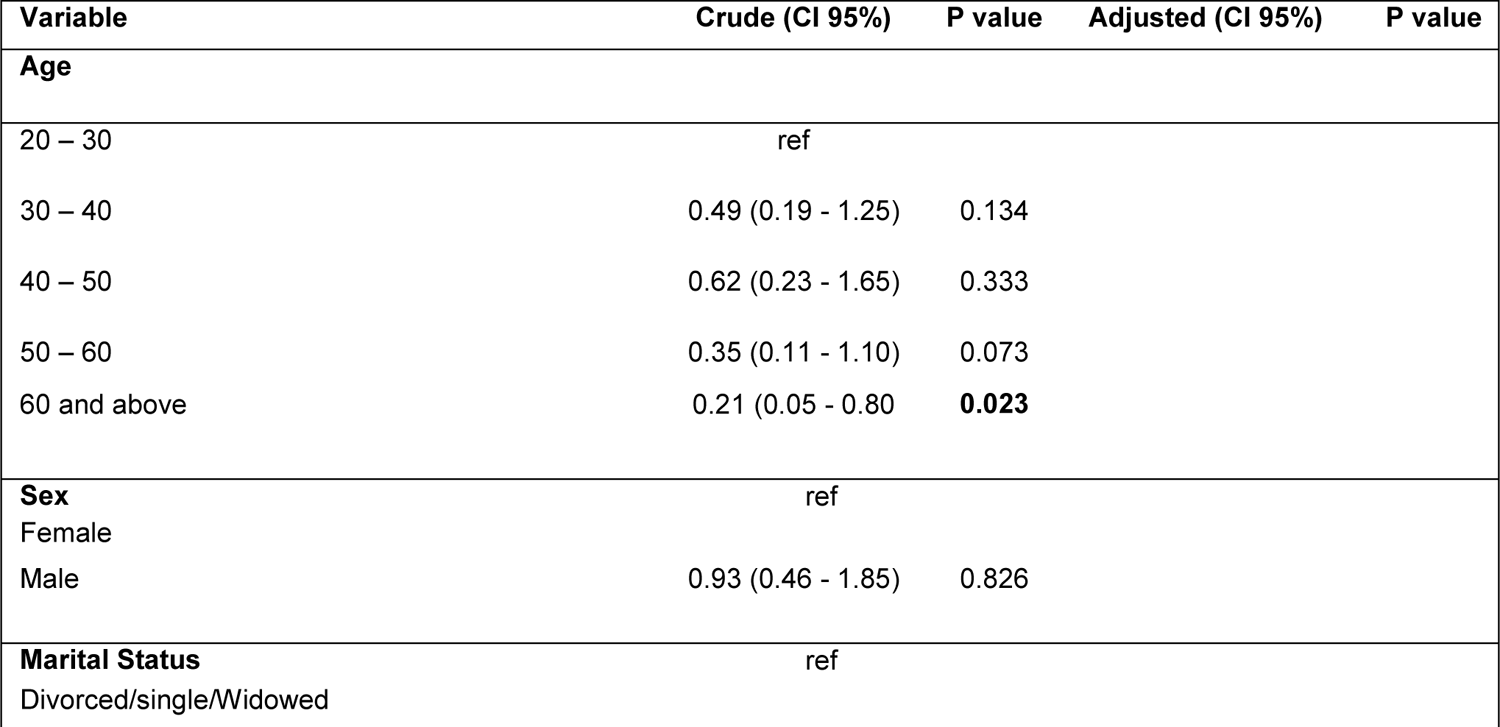
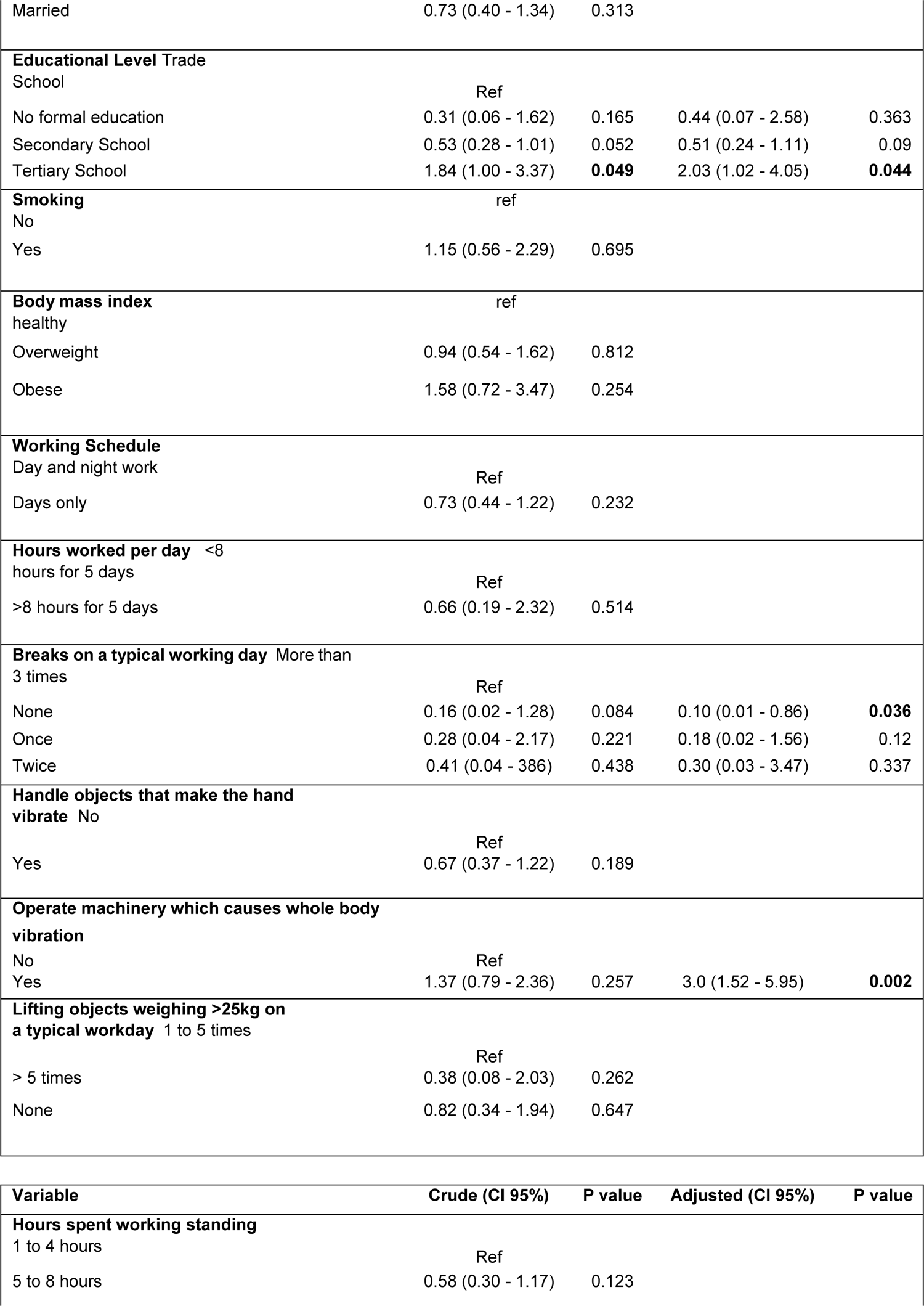
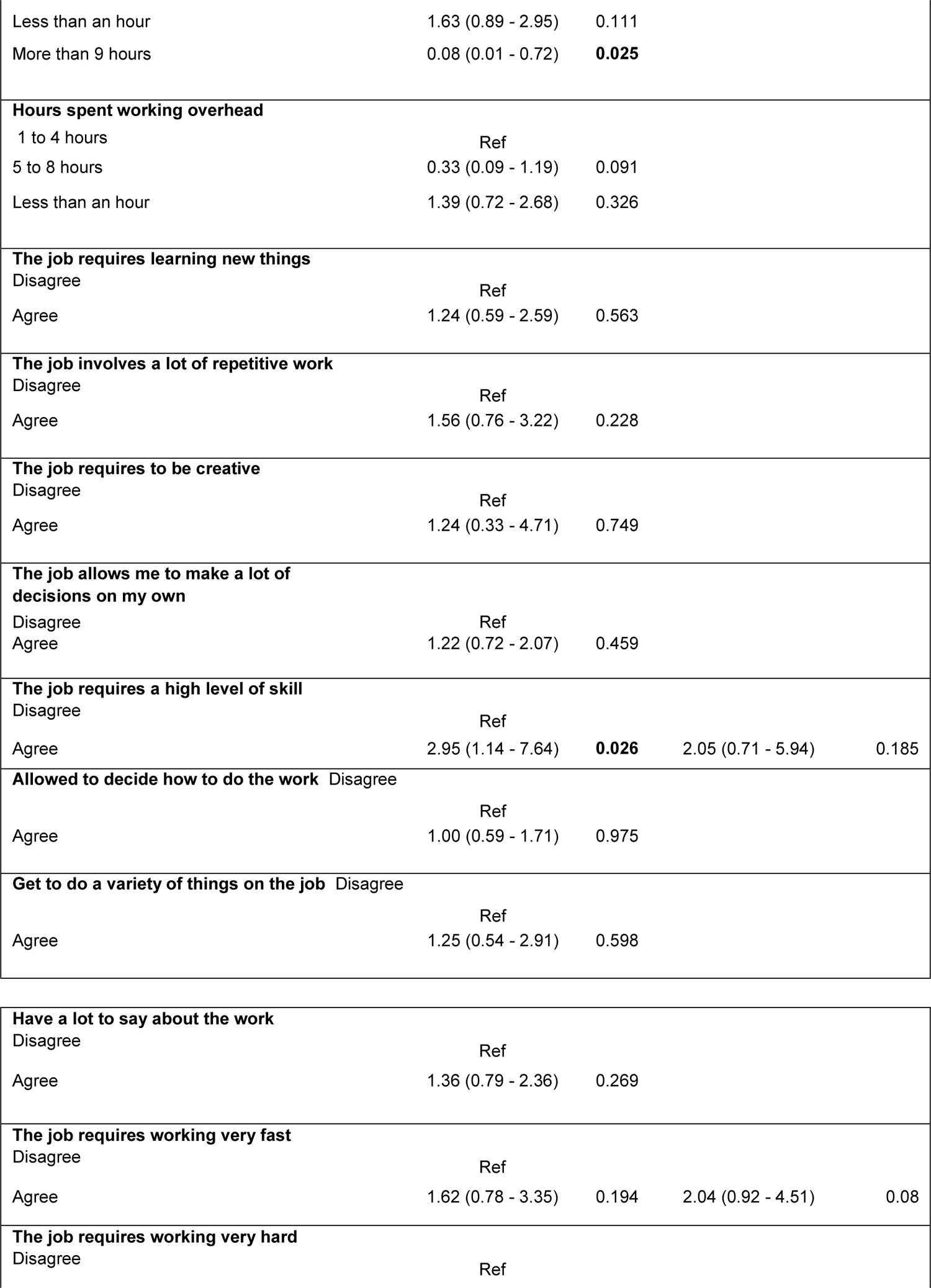
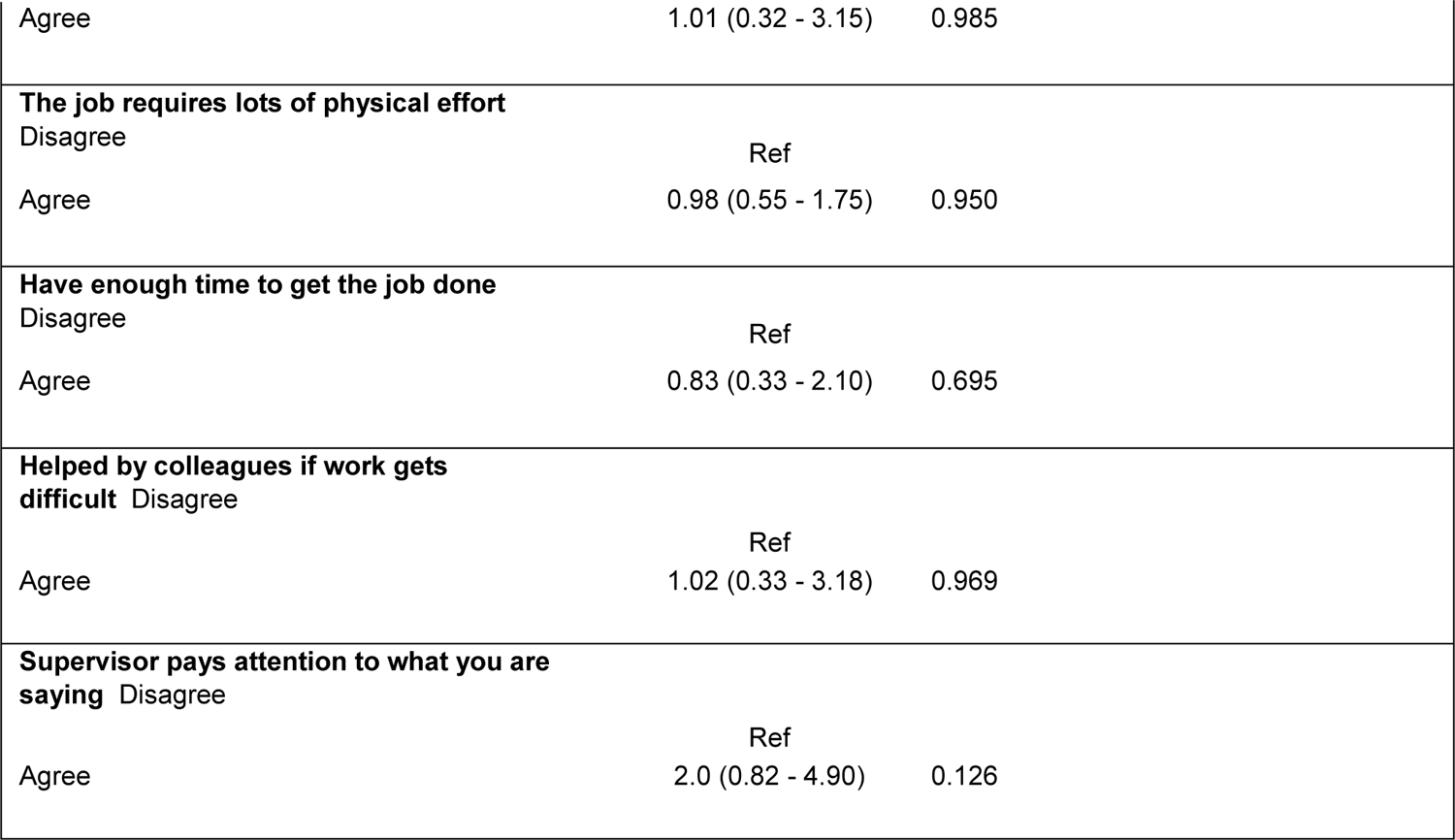
Factors associated with WMSDs among mineworkers at Kalumbila mine, Kalumbila District.

## Discussion

The study found that 77% of the participants had pain in at least one part of their body. The highest levels of discomfort or pain were reported in the lower back, upper back, and neck.

Associated with work-related musculoskeletal disorders in miners were participants with secondary education, no breaks during a typical working day, and operating machinery that produces whole-body vibration. The study’s findings confirm previous observations from multiple studies that suggest miners are associated with an increased prevalence of WMSD (10); (11); (12). The reported prevalence in this study is much higher when compared with that reported in previous studies conducted in China among construction workers (57.9%) (13), in Pakistan among construction workers (52%) (14), in Democratic republic of Congo among gold mine workers (61.2%) (5), and in India among surface miners (44.2%) (15). On the other hand, a much higher prevalence than that reported in this study is reported in Ghana among workers in the gold mining industry (85.5%) (11).

The most common area of pain was the lower back, followed by upper back and neck. These findings support findings from previous studies suggesting that lower back pain is the most common problem among miners (15); (14); (5); (11). Lower back pain is associated with the number of hours worked in a shift and the repetitive movements of a body part (16). This study reports a prevalence of 51.5%, which is consistent with the findings of (12), who reported a prevalence of 50.7%, but higher than that reported in the study from the Democratic Republic of Congo (14.8%) (5). Mine workers with a secondary education were found to be at a higher risk of developing WMSD than those with a vocational education.

The reasons for observing such results are unknown, however, there is some logical sense in suggesting that miners with secondary education may be more involved in manual work than miners with vocational training, which could put them at increased risk. This claim could be supported by the findings of a study which showed that manual mine workers were more susceptible to WMSDs than mechanised mine workers (17).

Socio-demographic factors such age, sex, length of service, body mass index and smoking were not associated with WMSD in this study. This is in contrast to the findings of previous studies which found that age (10); (13); (18), sex (18), body mass index (19), length of service (13); (18); (12) and smoking were associated with MSDs in workers (18). Work involving repetitive movements was not associated with WMSD in this study, which is contrary to previous studies that found repetitive movements to be associated with WMSD (12).

Working shifts were not associated with WMSDs, which conflicts with the findings of previous study which found that workers working two shifts had an increased risk of experiencing MSDs than those working a fixed day shift (18). Considering that only a few factors were associated with WMSDs in this study, there is still need for further studies to provide better understanding.

## Conclusion

The prevalence of WMSDs is relatively high among mine workers with more than three quarters experiencing pain/discomfort in at least one part of the body. Lower back disorders were the most common WMSDs among miners, followed by upper back and neck disorders, and these disorders were also found to cause the most work disability. These findings underscore the importance of investing and strengthening worker safety through ergonomic programs and workers’ training on safety measures to reduce the burden of work-related musculoskeletal disorders among miners. In addition, great attention should be paid to the factors associated with WMSD among miners, including those with secondary education and those who operate vibrating machinery.

## Data Availability

All data is available on request from the author

## Acknowledgements

I am most grateful to God Almighty for giving me the strength and protection to carry out this study. I wish to express my sincere gratitude to my Supervisor, Dr. Chisala Meki of the Department of Health Sciences, School of Public Health, UNZA for her patience, encouragement, corrections, and devoted time and energy in the preparation of this dissertation. I deeply recognize and appreciate the immense contributions of all other lecturers in the school for helping us navigate through the research. I would also like to thank my family for their support and encouragement to pursue this MPH program. Furthermore, God richly bless all those who have, in one way or the other, contributed to the successful write-up of this dissertation, especially my course mates and colleagues. Finally, I thank the team and research participants at Kalumbila Mine for their time and cooperation.

## Notes

### Competing Interest Statement

The authors have declared no competing interest.

### Funding Statement

The authors received no funding for this work

### Author Declarations

Ethics approval was obtained from the Ethics Review Committee through the School of Public Health at The University of Zambia. Consent was obtained from the biomedical research ethics committee and the National Health Regulatory Authority.

